# Longitudinal intergenerational patterns of psychopathology in a population-based dataset: an ABCD study

**DOI:** 10.64898/2026.09.04.26362261

**Authors:** Hajer Nakua, Jingxi Wang, Jennifer Warmingham, Nicole Racine, Marco Solmi, Rachel Marsh, Seonjoo Lee

## Abstract

**Objective:** To identify longitudinal trajectories of clinically relevant psychopathology from middle childhood to early adolescence and to examine the extent to which parental psychopathology influences child psychopathology trajectories.

**Method:** Participants were children and biological parents enrolled in the Adolescent Brain Cognitive Development (ABCD) Study (N=11,142) with up to five annual assessments of psychopathology using the Child Behavior Checklist and the Adult Self Report. Sequential latent class analysis identified longitudinal trajectories based on clinically elevated subscale scores (T-score≥65). Cross-lagged panel models examined bidirectional associations between parent and child internalizing and externalizing symptoms across development, including separate analyses of mother-child and father-child dyads.

**Results:** The majority of the sample (76.1%) showed stable psychopathology trajectories (67.9% low, 6.8% moderate, 1.3% high) of clinically elevated symptom endorsement. Nearly one-quarter of participants (21.9%) transitioned between trajectory classes over time (9.9% improved, 8.1% worsened, 5.9% fluctuating). Greater parental psychopathology was associated with increased odds of membership in the moderate- and high-symptom trajectory classes (ORs=1.28-2.61, p<0.001). Among 10,633 biological parent-child dyads, maternal internalizing symptoms demonstrated the strongest longitudinal associations with child internalizing symptoms 2-years later (standardized cross-lagged path β = 0.128, 95% CI = 0.094-0.161, p<0.001). Findings remained robust across sensitivity analyses.

**Conclusion:** These findings provide novel insight into population-level trends of psychopathology trajectories and the importance of understanding how parental psychopathology influences child psychopathology over the course of development. This study highlights the importance of maternal psychopathology as a risk factor for child psychopathology elevations across development.

## Introduction

Childhood and adolescence are characterized by substantial neurodevelopmental, cognitive, and social change, contributing to variability in the emergence and course of psychiatric symptoms^1–4^. Although the majority of children and youth exhibit relatively low and stable levels of psychopathology throughout development, there remains heterogeneity in the presentation and comorbidity of clinical levels of psychopathology symptoms^5,6^. Longitudinal studies of psychopathology trajectories can promote early identification of youth at elevated risk for psychiatric disorders prior to onset of a diagnosis. Longitudinal studies have typically employed variable-centered approaches, which estimate average developmental change within a population^7,8^, which provide insight into age-related trends in psychiatric symptoms. However, population-level trajectories may obscure meaningful individual-level variation in symptom development. Person-centered approaches address this limitation by identifying subgroups of individuals who follow unique developmental pathways, thereby providing insight into the prevalence and stability of distinct risk trajectories over time. Previous studies using person-centered approaches involved analyses using dimensional (continuous) symptom scores^6^. Characterizing developmental trajectories using clinically meaningful thresholds may provide insight into the proportion of youth who are at risk for a psychiatric condition. Once distinct developmental trajectories have been identified, examining risk factors associated with trajectory membership can provide insight into the heterogeneity of psychopathology outcomes.

Parental psychopathology is among the most robust risk factors of child psychopathology^9–11^. This *intergenerational transmission* reflects the extent to which psychiatric symptoms are present in consecutive generations. As such, parental psychopathology may be an important risk marker for child psychopathology presentation, either through predicting child membership within distinct developmental trajectories or predicting group-level variation in specific psychopathology domains in children. Parental psychopathology can inform the *type* of intergenerational transmission most likely to affect their children. *Homotypic transmission* refers to associations between the same symptom domains in parents and children, whereas *heterotypic transmission* refers to associations across different symptom domains^12–14^. Evidence supports both forms of transmission^12–14^, suggesting that parental psychopathology may confer broad vulnerability to child psychopathology. Despite extensive evidence linking parental and child psychopathology, several important gaps remain. First, the literature on intergenerational psychopathology has historically focused on maternal psychopathology^15–17^, limiting understanding of paternal contributions to child psychopathology. Second, many studies have examined parent-child psychopathology associations using cross-sectional designs^14,18,19^, providing limited insight into how these associations change throughout development at the population-level. Although longitudinal studies have examined associations between parental and child psychopathology, many have relied on small-to-moderate samples and have focused on associations between symptom domains rather than developmental patterns of psychopathology^11,13,20^. Finally, limited work has investigated whether parental psychopathology is associated with and can differentiate children who follow distinct developmental trajectories. As such, parental psychopathology symptoms can inform broad child psychopathology trajectories as well as the psychopathology domains most likely to affect their children. Evaluating the extent to which parental psychopathology predicts longitudinal child psychopathology presentation will provide insight into the pathways of influence of parental psychopathology on child outcomes; thereby promoting early identification of children with elevated psychopathology from high-risk families.

To address these gaps, the present study leverages longitudinal data from the Adolescent Brain Cognitive Development (ABCD) Study to expand on and integrate prior work examining both longitudinal psychopathology trajectories in children^6,8^ and parent-child psychopathology associations^11,21^. In Aim 1, we identify person-centered trajectories of clinically-relevant psychopathology symptoms across five annual assessments spanning middle childhood to early adolescence. We also test whether parent psychopathology predicts trajectory membership. In Aim 2, we examine longitudinal associations between parental and child psychopathology to characterize both homotypic and heterotypic transmission across development while considering unique contributions from both mothers and fathers. This study aims to clarify the prevalence and stability of clinically meaningful risk profiles and elucidate how parental psychopathology influences developmental patterns of pediatric psychopathology risk.

## Methods

### Sample

The ABCD dataset is a longitudinal multi-site community-derived, population-based sample collecting a comprehensive measurement battery (including genetic, blood, environmental, cognitive, brain, and behavioural measures) in > 11,000 9-11 year-old participants, with data collection time points occurring annually or biannually for 10 years. Participants were recruited from 21 academic sites across the U.S. using probability sampling to ensure that demographic trends across the U.S. are well represented in the sample^22^. Recruitment occurred through presentations and emails delivered to parents of children in local schools around each site. Interested parents underwent a telephone screening to determine whether their children were eligible to participate in the study. Participants were excluded from the ABCD study if they had MRI contraindications, no English fluency, uncorrected vision, hearing or impairments, major neurological disorders, were born extremely preterm (less than 28 weeks gestation), low birth weight (< 1200 grams), birth complications, or unwillingness to complete assessments. For Aim 1, we used child psychopathology tabulated data from six timepoints provided by the ABCD consortium (ABCD-6.1; n=11,142). Participants with missing psychopathology data in more than 3 timepoints were excluded from this analysis (see *Figure S1* for consort diagram and details of exclusion). We included multiple siblings in the main analysis of the study (see Supplementary Section 1). For Aim 2, we used parent and child psychopathology data from children and their biological parents (N=10,633).

### Child and Parent Psychopathology

Child psychopathology was indexed by the parent-reported Child Behavioral Checklist (CBCL, 6-18 years^23^). To address Aim 1, we used the 8 syndrome subscale T-scores from the parent-report CBCL for 6-18 year-olds: anxious/depressed, withdrawn/depressed, somatic complaints, thought problems, social problems, rule-breaking behaviour, aggressive behaviour, attention problems. To address Aim 2, broadband externalizing and internalizing symptom raw scores were used. See details in Supplementary Section 1.2.

Parent psychopathology was indexed by the Adult Self-Report (ASR; ages 18–59 years ^24^). Across both aims, we used the ASR broadband internalizing and externalizing symptom raw scores. We used the raw (unadjusted) score from the ASR as T-scores are not provided in the ABCD tabular data and there is no well-established T-score calculation for the ASR. See details in Supplementary Section 1.2.

### Statistical Analysis

#### Aim 1: Person-centered psychopathology trajectories across development

CBCL subscale T-scores were first dichotomized into clinically significant symptom endorsement; T-score ≥ 65 (1) and T-score < 65 (0). The dichotomization was used to identify clinically relevant trajectories. Sequential latent class analysis (SLCA) was conducted using the *slca* package in R. The SLCA identifies unobserved (latent) subgroups based on similar patterns of ordinal or categorical variables over time^25^. As such, the SLCA predicts how membership in one subgroup at one timepoint predicts the same subgroup membership at a different timepoint. The final solution of class number was selected based on AIC, BIC, class distribution, and descriptive assessments of class transition patterns. A first-order sequential transition structure was specified across adjacent longitudinal assessment waves, such that latent class membership at each assessment wave depended only on membership from the preceding wave (e.g., children who endorsed clinically significant symptoms at year 2 but not baseline would be considered to have changed classes). Model estimation was performed using the expectation-maximization (EM) algorithm implemented within the *estimate()* function in *slca* which provides a probability metric for how likely a given individual will belong to a subgroup. Missing data were handled in two steps. Prior to model estimation, participants were retained in the analytic sample if they had sufficient non-missing dichotomized CBCL syndrome subscale data, defined as data available across at least three assessment waves. Within the SLCA model, remaining partially missing responses were handled using full information maximum likelihood (FIML) under a Missing At Random (MAR) assumption. After model estimation, posterior marginal class probabilities were extracted for each wave. Individuals were assigned to the latent class with highest posterior probability at each wave. These wave-specific class assignments were then used to summarize longitudinal transition patterns and visualize transitions across waves. Individuals were assigned to the latent class with the highest posterior probability. Interpretation of longitudinal subgroup membership was facilitated through visualization of model-estimated and empirical transition probability matrices using heatmaps, alluvial flow diagrams, and longitudinal state-transition network graphs. We did not include covariates when estimating trajectories via SLCA for two reasons: 1) we aim to identify psychopathology trajectory groups that represent the ABCD sample and adding covariates may remove meaningful variance across trajectories; 2) given the dichotomous nature of the psychopathology input data, adding covariates would distort the interpretation of the models. We tested whether parental ASR scores would differ between the child symptom classes using an ANOVA (*Table S1*). We tested whether parental psychopathology symptoms, sex, child age at baseline and parental education increase the odds of children endorsing clinically meaningful psychopathology trajectories using multinomial logistic regression (*Table S2; S3*). We also tested whether retaining only one sibling per family would influence trajectory membership (*Table S4*).

#### Aim 2: Dynamic parent-child psychopathology relationships across time

We first calculated zero-order Pearson correlations between parent and child psychopathology subscale scores across three timepoints (baseline, year 2, year 4). We additionally calculated the zero-order Pearson correlations when stratifying the sample by parent and child sex.

We implemented a cross-lagged panel model (CLPM) analysis to evaluate longitudinal bidirectional associations between parent and child psychopathology across the baseline, 2-year, and 4-year timepoints using the *lavaan* package in R. We used CBCL and ASR internalizing and externalizing unadjusted (raw) broadband scores for this analysis. Separate models were fitted for either child internalizing and externalizing as the main outcome variable. Each model included child psychopathology (internalizing or externalizing), parent internalizing symptoms, and parent externalizing symptoms within a three-factor framework. Models included autoregressive paths, cross-lagged paths, and within-wave correlations across baseline, 2-year, and 4-year assessments. The standardized beta estimates of the cross-lagged paths were used to quantify the effect size of bidirectional parent-child associations over time. Child sex, child age at baseline, and parental education were included as covariates in all models, with covariate effects constrained to be equal across follow-up timepoints. Missing data were handled using FIML. Standard errors and confidence intervals were estimated using bootstrap resampling using the *sem* (se=bootstrap) function in the *lavaan* package with 5000 iterations. We tested whether retaining only one sibling per family would influence parent-child associations (*Table S5*). Given recent recommendations regarding the use of random-intercept cross-lagged panel models (RI-CLPM) to distinguish between-person from within-person effects^26,27^, we conducted sensitivity analyses comparing parent-child associations estimated using CLPM and RI-CLPM (*Table S6*).

*Reproducibility of parent-child associations:* To evaluate the robustness and reproducibility of the standard beta estimates of the CLPM, split-half analyses were conducted by randomly splitting the sample in half in 5000 different iterations (similar to prior work^28,29^). Within each of the 5000 iterations, we split the sample into two split-halves (A and B), and calculated a model-wide Pearson correlation coefficient encompassing the correlations across all the respective standardized beta estimates of each path between the two split-halves. We then calculated the contribution of each path in this Pearson correlation coefficient (1 coefficient for each iteration). For each pathway, the Z-score is the absolute mean pathway-specific contribution divided by the standard deviation of that same pathway-specific contribution across the 5,000 iterations. Larger Z-scores indicate a more reproducible contribution of that pathway to the model-wide split-half Pearson correlation. A reproducible distribution was defined as a Z-score magnitude > 1.96 which is associated with a *p* value < .05, indicating that the distribution significantly differed from zero, consistent with prior work^28,29^.

*Stratified Analysis:* To explore potential differences between mother-child and father-child psychopathology associations, we performed additional CLPM analyses stratifying for mother-child and father-child bidirectional associations. Caregiver subgroup classification was based on baseline caregiver identity variables derived from ABCD demographic assessments (92.5% remained stable across all three timepoints). We conducted a supplementary analysis with the caregivers who remained stable across all three timepoints to determine if results were consistent across time (*Table S7*). Standardized cross-lagged parameter estimates were descriptively compared across mother-only and father-only subgroup models to evaluate potential differences in homotypic and heterotypic parent-child psychopathology associations over time. Given the sample size difference between biological mothers (89.5%) and fathers (10.5%) in the current study, we performed a stabilized inverse probability weighting to evaluate whether parent-specific findings could be explained by differences in sample size and composition between the mother and father groups. Parent-specific propensity scores were estimated using logistic regression, with parent (mother or father) as the outcome and child sex, baseline child age, race, ethnicity, household income, and parental education as predictors. Stabilized weights of the ASR internalizing and externalizing scores for mothers and fathers were calculated from the estimated propensity scores. These weighted ASR scores were then inputted into an additional CLPM sensitivity analysis. Weighted CLPM estimates were then compared with the primary unweighted estimates to evaluate robustness and consistency of parent-specific models (*Table S8*).

### Code availability

The code for all analyses is available in a public GitHub repository (https://github.com/LunaJW/intergenerational-psychopathology).

## Results

The sample included in the current study was overrepresented by White American participants and higher parental education. See Table 1 for demographic details.

**Table 1.** Demographic and baseline clinical characteristics of the current sample.

| **Variable** | **Overall N = 11,142<sup>1</sup>** | **Mother caregiver**<br>**N = 9,525<sup>1</sup>** | **Father caregiver**<br>**N = 1,108<sup>1</sup>** |
| --- | --- | --- | --- |
| **Child age at baseline** | 9.96 (0.62) | 9.96 (0.63) | 9.94 (0.62) |
| **Child sex** |  |  |  |
| Male | 5,850 (52.5%) | 4,963 (52.1%) | 645 (58.2%) |
| Female | 5,292 (47.5%) | 4,562 (47.9%) | 463 (41.8%) |
| **Race** |  |  |  |
| White | 8,366 (75.1%) | 7,200 (75.6%) | 879 (79.3%) |
| Black | 1,785 (16.0%) | 1,545 (16.2%) | 98 (8.8%) |
| Asian | 280 (2.5%) | 179 (1.9%) | 62 (5.6%) |
| Other / Multiracial | 688 (6.2%) | 580 (6.1%) | 68 (6.1%) |
| Missing / Unknown | 23 (0.2%) | 21 (0.2%) | 1 (0.1%) |
| **Ethnicity** |  |  |  |
| Not Hispanic/Latino | 8,789 (78.9%) | 7,471 (78.4%) | 893 (80.6%) |
| Hispanic/Latino | 2,216 (19.9%) | 1,943 (20.4%) | 197 (17.8%) |
| Missing / Unknown | 137 (1.2%) | 111 (1.2%) | 18 (1.6%) |
| **Household income** |  |  |  |
| <\$25k | 1,448 (13.0%) | 1,302 (13.7%) | 72 (6.5%) |
| \$25k–\$49,999 | 1,480 (13.3%) | 1,284 (13.5%) | 117 (10.6%) |
| \$50k–\$99,999 | 2,915 (26.2%) | 2,466 (25.9%) | 309 (27.9%) |
| ≥\$100k | 4,403 (39.5%) | 3,668 (38.5%) | 558 (50.4%) |
| Missing / Declined | 896 (8.0%) | 805 (8.5%) | 52 (4.7%) |
| **Parental education** |  |  |  |
| Bachelor's degree or higher | 6,089 (54.6%) | 5,130 (53.9%) | 695 (62.7%) |
| Some college / Associate | 3,217 (28.9%) | 2,802 (29.4%) | 270 (24.4%) |
| High school or less | 1,821 (16.3%) | 1,580 (16.6%) | 141 (12.7%) |
| Missing / Declined | 15 (0.1%) | 13 (0.1%) | 2 (0.2%) |
| **Child CBCL internalizing score** | 53.94 (4.73) | 53.99 (4.76) | 53.04 (3.89) |
| **Child CBCL externalizing score** | 52.75 (4.79) | 52.70 (4.70) | 51.94 (3.60) |
| **Parent ASR internalizing score** | 9.49 (8.68) | 9.63 (8.70) | 8.14 (7.93) |
| **Parent ASR externalizing score** | 4.49 (4.79) | 4.48 (4.79) | 4.57 (4.76) |

<sup>1</sup>Values are mean (SD) for continuous variables and n (%) for categorical variables. The Overall column reflects the sample size used for Aim 1. Mother and father caregiver columns reflect the sample size(s) used for Aim 2; complete parent-child CBCL and ASR data of biological parents. Child CBCL scores are reported as mean T-score composites. Parent ASR scores are reported as raw sum scores.

**Table 2.** Mother-child and father-child cross-lagged panel model results.

| Outcome | Pathway | Mother ($\beta$, [95% CI], p value) | Father ($\beta$, [95% CI], p value) |
| --- | --- | --- | --- |
| Child internalizing model | Cross-lagged path: Parent internalizing baseline → Child internalizing 2-year | 0.128 [0.094, 0.161], p = <0.001 | 0.057 [-0.031, 0.145], p = 0.208 |
| Child internalizing model | Cross-lagged path: Parent externalizing baseline → Child internalizing 2-year | 0.031 [-0.003, 0.065], p = 0.073 | 0.056 [-0.037, 0.150], p = 0.236 |
| Child internalizing model | Cross-lagged path: Parent internalizing 2-year → Child internalizing 4-year | 0.112 [0.081, 0.143], p = <0.001 | 0.079 [-0.008, 0.167], p = 0.075 |
| Child internalizing model | Cross-lagged path: Parent externalizing 2-year → Child internalizing 4-year | 0.037 [0.005, 0.070], p = 0.022 | -0.015 [-0.098, 0.067], p = 0.718 |
| Child internalizing model | Autoregressive path: Child internalizing baseline → Parent internalizing 2-year | 0.036 [0.013, 0.059], p = 0.002 | 0.042 [-0.028, 0.113], p = 0.235 |
| Child internalizing model | Autoregressive path: Child internalizing 2-year → Parent internalizing 4-year | 0.049 [0.024, 0.074], p = <0.001 | 0.023 [-0.059, 0.104], p = 0.585 |
| Child internalizing model | Autoregressive path: Child internalizing baseline → Parent externalizing 2-year | 0.028 [0.004, 0.053], p = 0.022 | 0.033 [-0.047, 0.112], p = 0.417 |
| Child internalizing model | Autoregressive path: Child internalizing 2-year → Parent externalizing 4-year | 0.034 [0.010, 0.059], p = 0.006 | -0.008 [-0.094, 0.078], p = 0.855 |
| Child externalizing model | Cross-lagged path: Parent internalizing baseline → Child externalizing 2-year | 0.048 [0.012, 0.084], p = 0.009 | -0.001 [-0.087, 0.084], p = 0.976 |
| Child externalizing model | Cross-lagged path: Parent externalizing baseline → Child externalizing 2-year | 0.050 [0.011, 0.089], p = 0.011 | -0.004 [-0.097, 0.089], p = 0.932 |
| Child externalizing model | Cross-lagged path: Parent internalizing 2-year → Child externalizing 4-year | 0.031 [-0.006, 0.067], p = 0.098 | -0.010 [-0.102, 0.081], p = 0.822 |
| Child externalizing model | Cross-lagged path: Parent externalizing 2-year → Child externalizing 4-year | 0.088 [0.047, 0.129], p = <0.001 | -0.000 [-0.082, 0.082], p = 0.996 |
| Child externalizing model | Autoregressive path: Child externalizing baseline → Parent internalizing 2-year | -0.009 [-0.033, 0.014], p = 0.430 | 0.007 [-0.057, 0.070], p = 0.832 |
| Child externalizing model | Autoregressive path: Child externalizing 2-year → Parent internalizing 4-year | 0.020 [-0.007, 0.046], p = 0.151 | 0.002 [-0.092, 0.097], p = 0.961 |
| Child externalizing model | Autoregressive path: Child externalizing baseline → Parent externalizing 2-year | 0.014 [-0.013, 0.042], p = 0.296 | 0.034 [-0.057, 0.124], p = 0.466 |
| Child externalizing model | Autoregressive path: Child externalizing 2-year → Parent externalizing 4-year | 0.032 [0.003, 0.060], p = 0.029 | 0.001 [-0.100, 0.103], p = 0.981 |

Note: Standardized beta values in this table correspond to the specific path within the overall cross-lagged panel model.

### Aim 1: Person-centered psychopathology trajectories across development

The SLCA of the CBCL subscale scores at baseline identified a 3-class solution with the best model fit (log likelihood = -78916.86, AIC = 158185.7, BIC = 159473.8). These classes represent a low symptom group, a moderate symptom group, and high symptom group (see *Figure 1A; Table S5*). Overall, endorsement of various psychopathology domains was present in the moderate and high symptom group. The moderate symptom group featured children who endorsed a subset of clinically significant subscale scores. Among the high symptom group, the greatest endorsed psychopathology symptoms were thought problems. Children in the high symptom group were overrepresented by males, lower household income, and lower parental education (*Table S1*).

**Figure 1.**
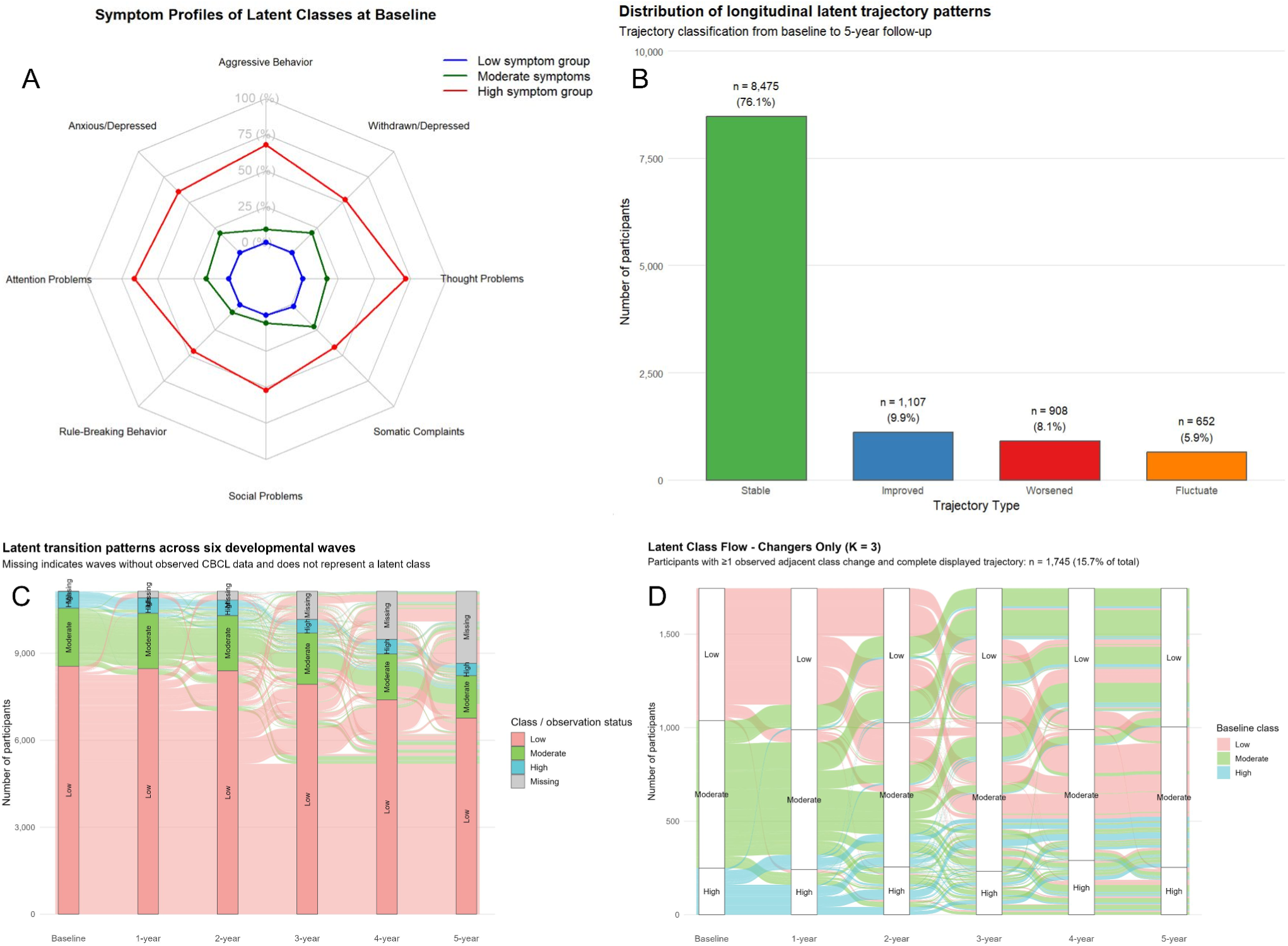
Sequential latent class analysis results using clinically meaningful cut-off scores in the ABCD sample. Note: (A) illustrates the radar plot describing the percentage of children in the low, moderate, and high group that endorsed the various CBCL subscales. An endorsement indicates that the T-score of a given subscale for a participant was >65 suggesting a clinically meaningful endorsement of these psychopathology traits. (B) Barplots depicting the number and percentage of participants that remained within their baseline assigned group (stable), improved over time, worsened over time, or fluctuated over time. (C) Latent transition patterns across the timepoints included in our study. (D) Flow chart depicting the children who changed classes across timepoints. While 2,439 children and youth in the current sample changed classes across time, this figure only depicts the subset that has complete data across all timepoints (n=1745).

**Figure 2.**
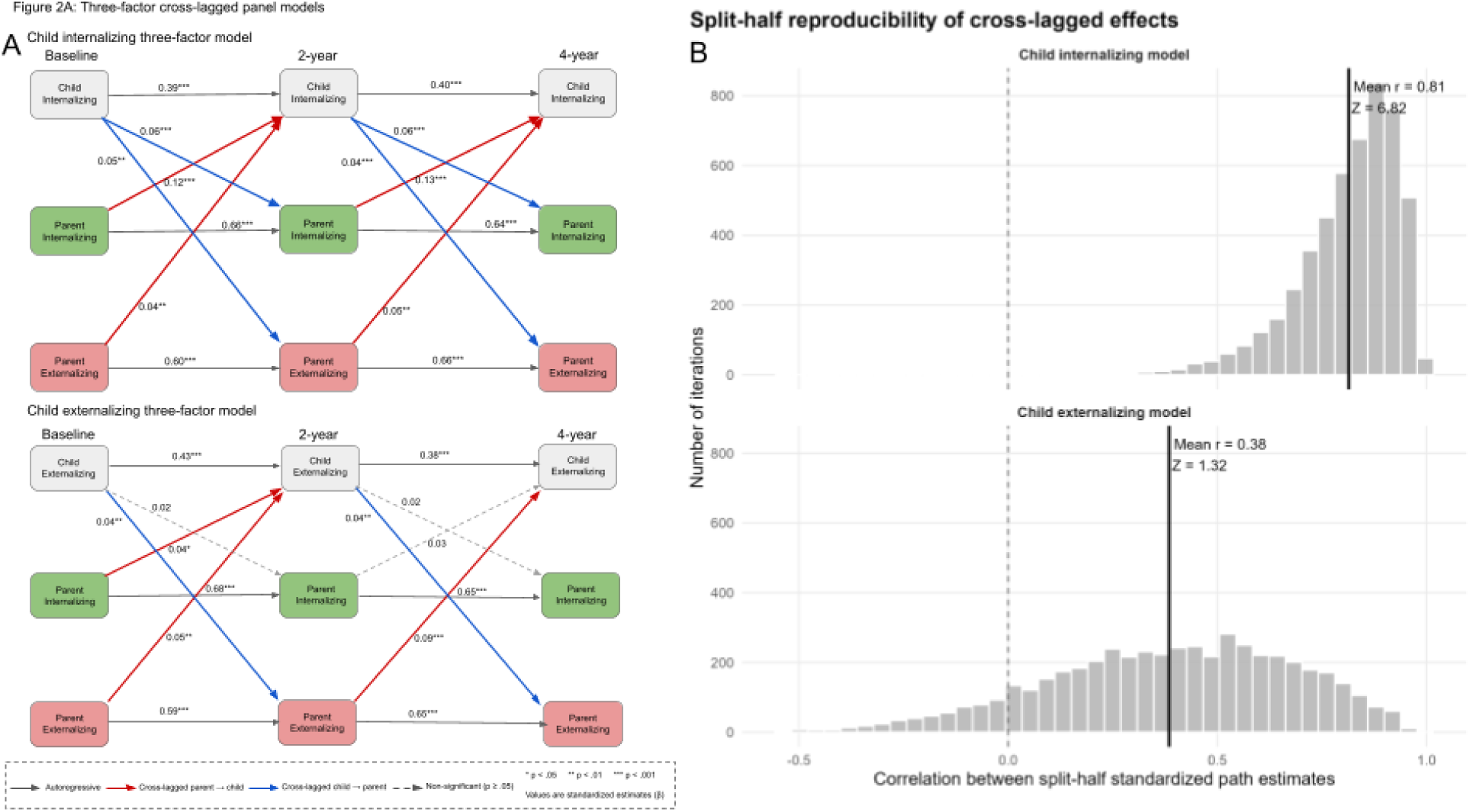
Cross-lagged panel models of parent-child bidirectional relationships in internalizing and externalizing symptoms. Note: (A) depicts cross-panel paths between parent-child and child-parent internalizing and externalizing symptoms as well as the autoregressive paths. Significant associations are noted by the standardized effect size with asterisks, and within-sample reproducible associations are depicted through the bolded path lines. (B) The results of the Pearson correlations (x-axis) of the standardized cross-lagged path estimates between the two split-halves across the 5000 iterations. A mean r-value closer to 1 indicates greater similarity in the standardized cross-lagged path estimates between split-halves, suggesting greater within-sample reproducibility. A Z-score >1.96 of a distribution is considered reproducible.

The SLCA revealed that 67.9% (n=7570) of children in the sample remained in the low symptom group across all time points, whereas 6.8% (n=758) and 1.3% (n=147) of children in the sample remained in the moderate and high symptom groups, respectively. The remainder of the children in the sample (21.9%, n=2439) transitioned between the symptom groups across timepoints (*Figure 1B*). *Figure 1C* shows the proportion of individuals who transitioned to different symptom classes across time. Nearly 10% (n=1107) showed decreases in psychopathology symptoms over time (*improved*), 8.1% (n=908) showed increases in clinically significant psychopathology scores over time (*worsened*), and 5.9% (n=652) showed fluctuating scores over time (*fluctuating*). The *worsened symptom group* all went from the low symptom group to the moderate symptom group. The timepoint with the greatest number of transitions occurred from year 2 (ages 11-12) to year 3 (ages 12-13) in which several children and youth who started in the moderate symptom group transitioned to low or high symptom group (*Figure 1D; Table S10*). Importantly, parent (biological mother or father) baseline internalizing and externalizing symptoms were significantly higher in the high symptom group compared to the moderate and low symptom group (*Table S1*). Parent baseline internalizing and externalizing symptoms significantly predicted the odds of their child’s psychopathology trajectory (*Table S2; S3*). For example, 1 standard deviation increase in parental internalizing scores predicted a nearly two fold increase in the likelihood of a child being classified into the moderate symptom group compared to the low symptom group (OR=1.99; 95%CI=1.86-2.14, p<0.001).

### Aim 2: Dynamic parent-child psychopathology relationships across time

Biological mothers accounted for 85.9% of the baseline biological parent caregiver sample, amongst which, 92.5% remained the caregiver who assessed their child in years 2 and 4. Zero-order correlations revealed significantly moderate correlations between respective parent self-report ASR subscales and parent-reported CBCL subscale scores at baseline, year two, and year four (*Figure S2A*). Parent-child correlations were strongest between internalizing domains. When stratifying the sample by child sex we found similar parent-child correlations (*Figure S2B*). Parent internalizing symptoms at baseline were significantly associated with child internalizing symptoms at year 2 (Standardized cross-lagged path β=0.12, 95% CI=0.090-0.151, p>0.001). Specifically, a 1-unit increase in parent internalizing symptoms predicted a 0.12-unit increase in child internalizing symptoms two years later. Similar trends occurred between parent internalizing symptoms at year two associated with child internalizing symptoms at year 4 (*Table S11*). We also found significant child-to-parent relationships, however, these were smaller in magnitude than parent-to-child relationships (*Figure 3; Table S11;* equality-constraint tests presented in *Table S12*). For example, a 1-unit increase in child internalizing behaviour at baseline was associated with a 0.044-unit increase in parent internalizing behaviour two-years later (95% CI=0.022-0.066, p<0.001). Homotypic parent-child externalizing associations were significant, yet showed small effect sizes. Similarly, effect sizes were small for heterotypic parent-child psychopathology associations. Split-half resampling analyses revealed that the most reproducible effect was homotypic relationships between parent-to-child internalizing symptoms (Z-score = 6.82; mean r-value = 0.81; *Figure 3*).

**Figure 3.**
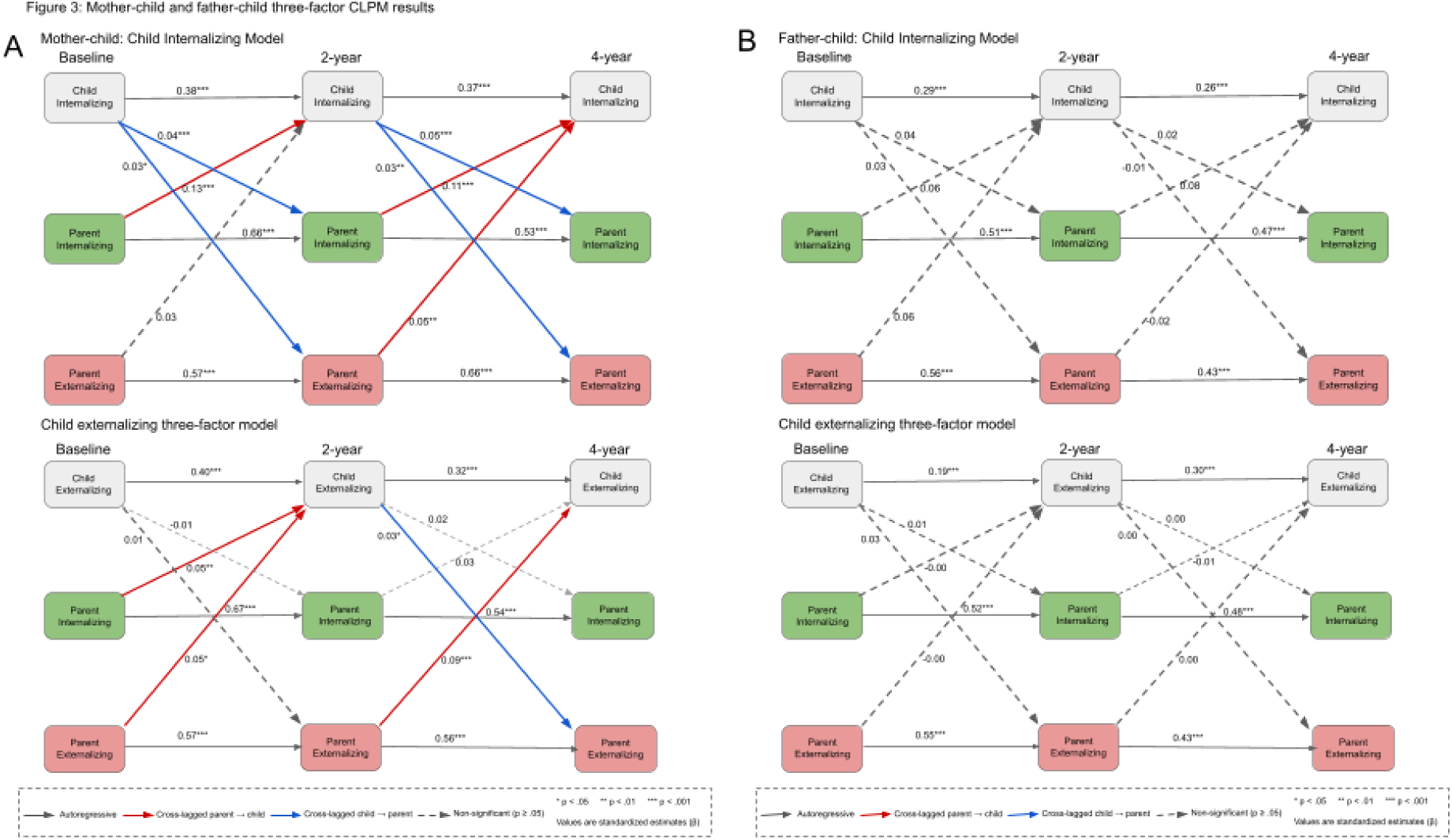
Cross-lagged panel models of mother-child and father-child bidirectional relationships in internalizing and externalizing symptoms. Note: (A) depicts cross-panel paths between mother-child and child-mother internalizing and externalizing symptoms as well as the autoregressive paths. Significant associations are noted by the standardized effect size with asterisks, and within-sample reproducible associations are depicted through the bolded path lines. (B) depicts cross-panel paths between father-child and child-father internalizing and externalizing symptoms as well as the autoregressive paths. Significant associations are noted by the standardized effect size with asterisks, and within-sample reproducible associations are depicted through the bolded path lines.

*Sensitivity Analysis:* Mother-to-child internalizing symptom homotypic relationships showed the largest and most reproducible effect across all models (β=0.128; 95% CI = 0.094-0.161; p<0.001; *Table 4; Figure 3*). Father-to-child homotypic relationships across internalizing and externalizing symptoms were significant but not reproducible (*Table 4; Figure 3*). Similar mother-to-child effect sizes and reproducibility were found from baseline-to-year 2 and year 2-to-year 4, suggesting that maternal influence on child psychopathology is consistent throughout mid-to-late childhood. Child-to-mother and child-to-father effects were significant, yet not reproducible (*Table S8*). When stratifying the sample based on caregivers who completed the ASR and CBCL at each time point, we find consistent results to the main findings of Aim 2 (*Table S7*). Propensity weighted mother and father ASR scores were relatively consistent with unweighted scores suggesting that these results were not driven by the bias of an overrepresented sample of mothers (*Table S8*).

## Discussion

The present study leveraged longitudinal data from the ABCD Study to 1) characterize clinically meaningful developmental trajectories of psychopathology across middle childhood to early adolescence, and, 2) examine the influence of parental psychopathology on child psychopathology. Three main findings emerged. First, three distinct stable childhood psychopathology trajectories emerged characterizing low, moderate, and high symptom presentation, and representing 76.1% of the sample. The remaining 23.9% of the sample was characterized by improved, worsened or fluctuating psychopathology symptoms over time.

Second, elevated parental psychopathology symptoms predicted children being classified in the higher symptom trajectory groups. Third, parental psychopathology symptoms at baseline were associated with elevated child psychopathology symptoms 2 years later. Maternal internalizing symptoms demonstrated the strongest and most reproducible longitudinal associations with child internalizing symptoms, supporting the importance of maternal mental health as a critical pathway towards improving child developmental outcomes.

Consistent with prior population-based psychopathology trajectories^6^, the majority of youth in the present study remained within a low-symptom trajectory across the five-year follow-up period, with a substantial minority showing stable moderate and high symptom trajectories. However, nearly one-quarter of youth had fluctuations, either for better or worse, between clinical presentations of psychopathology over time highlighting the dynamic nature of clinical symptom presentations during late childhood and early adolescence. Approximately 4.3% of the total sample fluctuated in their class membership between year 2 (ages 11-12) to year 3 (ages 12-13). This period may represent a developmentally vulnerable period of change in psychopathology symptoms, consistent with prior findings of hormonal, social and neurobiological changes during this time period^30,31^. More specifically, nearly 9% of the sample classified into the low symptom group at baseline showed increases in psychopathology symptoms and were classified into the moderate symptom group over time (*Table S9; S10*).

These findings suggest that a substantial minority of children experience worsening mental health during this developmental period. The prevalence of persistently elevated psychopathology in the current study (1.3% of the sample) was lower than estimates reported in a prior person-centered trajectory study^6^. One likely explanation is methodological. Previous studies often defined trajectory groups using continuous symptom distributions, however, the present study used clinical thresholds to characterize trajectories. As such, the persistently elevated group identified here may represent a more severe subset of youth exhibiting persistently high levels of psychopathology. Notably, none of the participants showed clinical worsening from the low-symptom group to the high-symptom group across timepoints. It is possible that clinically-relevant psychopathology may be established early in childhood and persist across development, or may present gradually over time and present clinically during adolescence^32–34^.

In the current study, we consistently found a broad co-occurrence of clinically-relevant symptom endorsements across psychopathology domains among youth in the moderate- and high-symptom groups. This pattern is consistent with transdiagnostic models emphasizing a general liability toward psychopathology (i.e., the p-factor and HiTOP frameworks)^35,36^.

Developmental trajectories of psychopathology may reflect differences in overall psychopathology burden rather than progression within specific domains. Importantly, while a broad endorsement was found, the moderate symptom group was largely characterized by the somatic complaints domain, which measures physical symptoms that are thought to reflect emotional distress^37,38^, such as headaches, nausea, and stomach aches. This is consistent with findings that somatic symptoms are an early indicator of psychopathology during childhood and adolescence^39^. Sex differences between trajectories revealed that males were overrepresented in the moderate and high symptom groups (*Tables S1;S2;S13;S14*), however females were overrepresented in the worsened group (*Tables S13; S14*). These findings point to sex-specific vulnerability trajectories of psychopathology during middle to late childhood. Males may experience earlier peaks in psychopathology symptoms that stabilize during middle to late childhood, whereas females may be more vulnerable to elevated psychopathology during specific developmental windows.

Homotypic transmission between maternal and child internalizing symptoms was the strongest and most reproducible longitudinal parent-child association observed in the present study. This finding is consistent with prior work demonstrating that maternal depression, anxiety, and related internalizing symptoms are associated with elevated internalizing symptoms in children^15,16,40^. Several mechanisms may contribute to this finding. Intergenerational transmission of internalizing psychopathology is thought to arise through a combination of genetic liability and phenotypic transmission^12,13,41^, such as parental modelling of emotional responses and behaviours^13^. Prior work has suggested that direct phenotypic transmission may account for unique variance in child psychopathology beyond direct genetic transmission alone^13,42^. Mothers across families report spending more time caregiving during childhood compared to fathers^43,44^, potentially creating more opportunities for children to observe and adopt maternal emotional responses and behaviours. In contrast, father-child associations were weaker than mother-child associations and not reproducible, albeit statistically significant. The direction of father-child effects were consistent with mother-child effects, as such, we warrant caution against interpreting these results as evidence for the absence of paternal influences on child psychopathology. There are several potential explanations for the weak and non-reproducible father-child associations, including true small effects of paternal influence on child psychopathology that require large samples to be robust, differences in caregiving involvement between mothers and fathers, and/or developmental differences in the timing of paternal influences on child psychopathology^21,43–46^. Evidence supporting homotypic transmission effects, i.e., shared parent-child relationships within the same domain, of internalizing symptoms was found, whereas evidence supporting externalizing or heterotypic transmission, i.e., shared parent-child relationships within different domains, was limited. The cross-sectional correlations (*Figure S2*) indicated that parent-child externalizing associations were strongest at baseline and weakened over time. Although speculative, it is possible that intergenerational influences on externalizing symptoms may decrease as children progress through adolescence.

There are several limitations to consider when interpreting the results of the current study. First, child psychopathology was assessed using parent-reported CBCL measures, introducing the possibility of shared method variance and reporter bias, i.e., parents with more symptoms also reporting more symptoms in their children; this bias is supported by prior investigations^15^. Although such reporting biases may partially contribute to observed parent-child associations, they are unlikely to fully account for the reproducible homotypic internalizing relationships observed across time points. Third, the use of clinically relevant subscale thresholds may reduce sensitivity to subthreshold symptom variation. Fourth, while the population-based design used in the current study provides insight into the general trends of children in the US, the ABCD study does not provide a full representation of all the demographic groups within the US^47,48^. Finally, we only included biological mothers and fathers in Aim 2. While the scope of the study was focused on biological parent-child dyads, we recognize that non-biological caregiver-child relationships may also show phenotypic transmission and are important to study.

## Conclusion

This study addresses several gaps in the literature surrounding parental psychopathology influences on child psychopathology. Person-centered trajectories were marked by broad psychopathology comorbidity, supporting transdiagnostic models of psychiatric risk. Our findings provide evidence to support elevated parental psychopathology as a candidate risk marker of clinically-relevant child psychopathology symptom trajectories and increased psychopathology symptoms at the group-level throughout middle to late childhood. The results of this study pave the way for future research to examine more specific mechanistic pathways that underlie parent-to-child intergenerational transmission of psychopathology. We recommend three future directions: 1) examining how parental psychopathology influences the interactions between child brain and psychopathological development, 2) incorporating early developmental periods into longitudinal studies, 3) incorporating larger paternal samples to clarify the role of intergenerational transmission of psychopathology.

## Data Availability

Data for the ABCD Study are available through the National Institutes of Health [NIH] Brain Development Cohort (NBDC) Data Hub (https://www.nbdc-datahub.org/abcd-study).

https://github.com/LunaJW/intergenerational-psychopathology

## Acknowledgements

Data used in the preparation of this article were obtained from the Adolescent Brain Cognitive Development (ABCD) Study (https://abcdstudy.org), held in the NIMH Data Archive (NDA). This is a multisite, longitudinal study designed to recruit more than 10,000 children ages 9–10 and follow them over 10 years into early adulthood. The ABCD Study is supported by the National Institutes of Health and additional federal partners under award numbers U01DA041048, U01DA050989, U01DA051016, U01DA041022, U01DA051018, U01DA051037, U01DA050987, U01DA041174, U01DA041106, U01DA041117, U01DA041028, U01DA041134, U01DA050988, U01DA051039, U01DA041156, U01DA041025, U01DA041120, U01DA051038, U01DA041148, U01DA041093, U01DA041089, U24DA041123, U24DA041147. A full list of supporters is available at https://abcdstudy.org/federal-partners.html. A listing of participating sites and a complete listing of the study investigators can be found at https://abcdstudy.org/consortiummembers/. ABCD consortium investigators designed and implemented the study and/or provided data but did not necessarily participate in analysis or writing of this report. This manuscript reflects the views of the authors and may not reflect the opinions or views of the NIH or ABCD consortium investigators. The ABCD data repository grows and changes over time. The ABCD data used in this report came from NDA Release 6.1 (https://doi.org/10.82525/ab4q-qr87).

