## Supplemental File for "Longitudinal intergenerational patterns of psychopathology in a population-based dataset: an ABCD study"

### 1. ABCD Sample Used

#### *1.1. Data from multiple siblings*

In the primary analyses for Aims 1 and 2, we retained all children that met our inclusion criteria, including siblings from the same family. The objective of Aim 1 was to characterize clinically meaningful CBCL-derived psychopathology trajectories among children in the ABCD cohort rather than estimate family-level effects. Analyzing all eligible children maximized statistical power and allowed characterization of the full range of observed developmental trajectories. Given that siblings share genetic and environmental influences, observations within families are not statistically independent. To evaluate the potential impact of this dependence, we repeated the analyses after restricting the sample to one child per family by retaining the first enrolled sibling and excluding additional siblings. We found that retaining one child per family did not meaningfully change the results of Aim 1 or Aim 2 (Tables S4; S5), suggesting that any between-sibling variation did not meaningfully affect longitudinal child psychopathology trajectories of parent-child psychopathology relationships.

#### *1.2. Details of psychopathology scales*

*Child Psychopathology*

The Child Behavioral Checklist (CBCL, 6-18 years) is a well-validated parent/caregiver-reported tool to assess mental health symptoms in children (Achenbach & Rescorla, 2001). The CBCL consists of 113 questions about a child's behaviour on an ordinal scale (0 = never, 1 = sometimes, 2 = often). The ordinal ratings are summed to provide subscale scores for a variety of behaviours (e.g., aggression). To address Aim 1, we used the 8 syndrome subscale T-scores from the parent-report CBCL for 6–18 year-olds: anxious/depressed, withdrawn/depressed, somatic complaints, thought problems, social problems, rule-breaking behaviour, aggressive behaviour, attention problems. Participants were stratified between those who had a T-score >65 (clinically significant cut off) and those who did not. To address Aim 2, broadband externalizing and internalizing symptom raw scores were used. The internalizing score is calculated by combining the withdrawn, anxious/depressed and somatic CBCL subscales. The externalizing score is calculated by combining rule-breaking and aggressive CBCL subscales.

*Parent Psychopathology*

The Adult Self-Report (ASR; ages 18–59 years) is a well-validated self-report measure of adult psychopathology developed as part of the Achenbach System of Empirically Based Assessment (ASEBA; Achenbach & Rescorla, 2003). The ASR consists of 126 problem items assessing emotional, behavioural, and social functioning over the previous six months. Respondents rate each item on an ordinal scale (0 = not true, 1 = somewhat or sometimes true, 2 = very true or often true). Item ratings are summed to generate syndrome subscale scores. To address Aim 2, we used the ASR broadband Internalizing and Externalizing Problems T-scores. The Internalizing Problems score is derived from the Anxious/Depressed, Withdrawn, and Somatic Complaints syndrome scales, whereas the Externalizing Problems score is derived from the Rule-Breaking Behavior and Aggressive Behavior syndrome scales. We used the raw (unadjusted) score from the ASR as T-scores are not provided in the ABCD tabular data and there is no well-established T-score calculation for the ASR.

### 2. Tables

**Table S1. Demographic and baseline clinical characteristics by baseline latent class in the LTA analytic sample**

| Variable | Overall N = 11,142[1] | Low N = 8,555[1] | Moderate N = 2,005[1] | High N = 582[1] | p-value[2] |
|---|---|---|---|---|---|
| **Child age at baseline** | 9.96 (0.62) | 9.96 (0.62) | 9.95 (0.62) | 9.96 (0.64) | 0.859 |
| **Child sex** | | | | | <0.001 |
| Male | 5,850 (52.5%) | 4,391 (51.3%) | 1,092 (54.5%) | 367 (63.1%) | |
| Female | 5,292 (47.5%) | 4,164 (48.7%) | 913 (45.5%) | 215 (36.9%) | |
| **Race** | | | | | <0.001 |
| White | 8,366 (75.1%) | 6,385 (74.6%) | 1,558 (77.7%) | 423 (72.7%) | |
| Black | 1,785 (16.0%) | 1,371 (16.0%) | 289 (14.4%) | 125 (21.5%) | |
| Asian | 280 (2.5%) | 248 (2.9%) | 29 (1.4%) | 3 (0.5%) | |
| Other / Multiracial | 688 (6.2%) | 533 (6.2%) | 125 (6.2%) | 30 (5.2%) | |
| Missing / Unknown | 23 (0.2%) | 18 (0.2%) | 4 (0.2%) | 1 (0.2%) | |
| **Ethnicity** | | | | | 0.822 |
| Not Hispanic/Latino | 8,789 (78.9%) | 6,742 (78.8%) | 1,588 (79.2%) | 459 (78.9%) | |
| Hispanic/Latino | 2,216 (19.9%) | 1,713 (20.0%) | 388 (19.4%) | 115 (19.8%) | |
| Missing / Unknown | 137 (1.2%) | 100 (1.2%) | 29 (1.4%) | 8 (1.4%) | |
| **Household income** | | | | | <0.001 |
| <$25k | 1,448 (13.0%) | 997 (11.7%) | 304 (15.2%) | 147 (25.3%) | |
| $25k–$49,999 | 1,480 (13.3%) | 1,058 (12.4%) | 308 (15.4%) | 114 (19.6%) | |
| $50k–$99,999 | 2,915 (26.2%) | 2,226 (26.0%) | 545 (27.2%) | 144 (24.7%) | |

| | | | | | |
|---|---|---|---|---|---|
| ≥$100k | 4,403 (39.5%) | 3,599 (42.1%) | 687 (34.3%) | 117 (20.1%) | |
| Missing / Declined | 896 (8.0%) | 675 (7.9%) | 161 (8.0%) | 60 (10.3%) | |
| **Parental education** | | | | | <0.001 |
| Bachelor's degree or higher | 6,089 (54.6%) | 4,852 (56.7%) | 1,027 (51.2%) | 210 (36.1%) | |
| Some college / Associate | 3,217 (28.9%) | 2,320 (27.1%) | 639 (31.9%) | 258 (44.3%) | |
| High school or less | 1,821 (16.3%) | 1,372 (16.0%) | 337 (16.8%) | 112 (19.2%) | |
| Missing / Declined | 15 (0.1%) | 11 (0.1%) | 2 (0.1%) | 2 (0.3%) | |
| **Child CBCL internalizing score** | 53.94 (4.73) | 52.28 (2.58) | 57.96 (4.86) | 64.62 (6.30) | <0.001 |
| **Child CBCL externalizing score** | 52.75 (4.79) | 51.26 (2.29) | 55.37 (5.00) | 65.61 (7.35) | <0.001 |
| **Parent ASR internalizing score** | 9.49 (8.68) | 7.53 (6.87) | 14.45 (9.67) | 21.21 (12.26) | <0.001 |
| **Parent ASR externalizing score** | 4.49 (4.79) | 3.55 (3.88) | 6.75 (5.37) | 10.52 (7.19) | <0.001 |

[1]Values are mean (SD) for continuous variables and n (%) for categorical variables. Child CBCL scores are reported as mean T-score composites. Parent ASR scores are reported as raw sum scores. [2]One-way analysis of means; Pearson's Chi-squared test; Pearson's Chi-squared test with simulated p-value (based on 10000 replicates)

**Table S2. Baseline parent psychopathology predicting baseline child latent class membership**

| Predictor | OR [95% CI] | p-value |
|---|---|---|
| **Moderate vs Low** | | |
| Intercept | 0.20 [0.09, 0.47] | <0.001 |
| Parent ASR internalizing, per 1 SD increase | 1.99 [1.86, 2.14] | <0.001 |
| Parent ASR externalizing, per 1 SD increase | 1.28 [1.20, 1.37] | <0.001 |

| | | |
|---|---|---|
| Child sex: Female vs Male | 0.86 [0.77, 0.95] | 0.004 |
| Child age at baseline | 1.01 [0.93, 1.10] | 0.735 |
| Household income: $25,000–$49,999 vs <$25,000 | 1.06 [0.87, 1.28] | 0.587 |
| Household income: $50,000–$99,999 vs <$25,000 | 0.98 [0.82, 1.16] | 0.811 |
| Household income: ≥$100,000 vs <$25,000 | 0.95 [0.80, 1.12] | 0.557 |
| Household income: Missing/Declined vs <$25,000 | 0.96 [0.76, 1.21] | 0.729 |
| Parental education | 1.00 [1.00, 1.00] | 0.669 |
| **High vs Low** | | |
| Intercept | 0.03 [0.01, 0.14] | <0.001 |
| Parent ASR internalizing, per 1 SD increase | 2.49 [2.25, 2.76] | <0.001 |
| Parent ASR externalizing, per 1 SD increase | 1.58 [1.44, 1.74] | <0.001 |
| Child sex: Female vs Male | 0.58 [0.48, 0.70] | <0.001 |
| Child age at baseline | 1.09 [0.94, 1.26] | 0.276 |
| Household income: $25,000–$49,999 vs <$25,000 | 0.90 [0.67, 1.21] | 0.475 |
| Household income: $50,000–$99,999 vs <$25,000 | 0.69 [0.53, 0.91] | 0.008 |
| Household income: ≥$100,000 vs <$25,000 | 0.50 [0.38, 0.67] | <0.001 |
| Household income: Missing/Declined vs <$25,000 | 0.93 [0.65, 1.32] | 0.684 |
| Parental education | 1.00 [1.00, 1.00] | 0.615 |
| **High vs Moderate** | | |
| Intercept | 0.16 [0.03, 0.73] | 0.018 |
| Parent ASR internalizing, per 1 SD increase | 1.25 [1.13, 1.38] | <0.001 |
| Parent ASR externalizing, per 1 SD increase | 1.23 [1.12, 1.36] | <0.001 |
| Child sex: Female vs Male | 0.67 [0.55, 0.82] | <0.001 |

| | | |
|---|---|---|
| Child age at baseline | 1.07 [0.92, 1.25] | 0.384 |
| Household income: $25,000–$49,999 vs <$25,000 | 0.85 [0.63, 1.15] | 0.292 |
| Household income: $50,000–$99,999 vs <$25,000 | 0.71 [0.53, 0.94] | 0.015 |
| Household income: ≥$100,000 vs <$25,000 | 0.53 [0.39, 0.71] | <0.001 |
| Household income: Missing/Declined vs <$25,000 | 0.97 [0.67, 1.40] | 0.863 |
| Parental education | 1.00 [1.00, 1.00] | 0.443 |

[1] Multinomial logistic regression was used. Parent ASR internalizing and externalizing scores were standardized, so odds ratios reflect the change in odds of class membership per 1-SD increase in parent symptoms. Models adjusted for child sex, child age at baseline, household income, and parental education.

**Table S3. Baseline parent psychopathology predicting baseline child latent class membership in the subset of children who changed class membership overtime**

| Predictor | OR [95% CI] | p-value |
|---|---|---|
| **Worsened vs Improved** | | |
| Parent ASR internalizing, per 1 SD increase | 0.82 [0.74, 0.92] | <0.001 |
| Parent ASR externalizing, per 1 SD increase | 0.92 [0.83, 1.03] | 0.135 |
| Child sex: Female vs Male | 3.00 [2.50, 3.61] | <0.001 |
| Child age at baseline | 0.90 [0.78, 1.04] | 0.155 |
| Household income: 25, 000–49,999 vs <$25,000 | 0.92 [0.66, 1.29] | 0.634 |
| Household income: 50, 000–99,999 vs <$25,000 | 1.17 [0.86, 1.59] | 0.329 |
| Household income: ≥100, 000*vs* <25,000 | 1.15 [0.83, 1.60] | 0.408 |

| | | |
|---|---|---|
| Household income: Missing/Declined vs <$25,000 | 0.74 [0.49, 1.11] | 0.143 |
| Parental education: Some college / Associate vs Bachelor's degree or higher | 0.82 [0.65, 1.03] | 0.091 |
| Parental education: High school or less vs Bachelor's degree or higher | 0.78 [0.58, 1.06] | 0.116 |
| **Fluctuate vs Improved** | | |
| Parent ASR internalizing, per 1 SD increase | 0.96 [0.86, 1.07] | 0.417 |
| Parent ASR externalizing, per 1 SD increase | 1.02 [0.91, 1.13] | 0.750 |
| Child sex: Female vs Male | 1.50 [1.24, 1.83] | <0.001 |
| Child age at baseline | 0.92 [0.78, 1.07] | 0.268 |
| Household income: 25, 000–49,999 vs <$25,000 | 1.03 [0.73, 1.44] | 0.886 |
| Household income: 50, 000–99,999 vs <$25,000 | 0.91 [0.66, 1.27] | 0.584 |
| Household income: ≥100, 000*vs* <25,000 | 1.03 [0.72, 1.46] | 0.887 |
| Household income: Missing/Declined vs <$25,000 | 1.13 [0.76, 1.67] | 0.539 |
| Parental education: Some college / Associate vs Bachelor's degree or higher | 0.90 [0.70, 1.16] | 0.425 |
| Parental education: High school or less vs Bachelor's degree or higher | 0.86 [0.62, 1.18] | 0.350 |
| **Fluctuate vs Worsened** | | |
| Parent ASR internalizing, per 1 SD increase | 1.16 [1.03, 1.31] | 0.017 |

| | | |
|---|---|---|
| Parent ASR externalizing, per 1 SD increase | 1.10 [0.98, 1.25] | 0.105 |
| Child sex: Female vs Male | 0.50 [0.41, 0.62] | <0.001 |
| Child age at baseline | 1.02 [0.86, 1.20] | 0.847 |
| Household income: $25,000$–49,999 vs <$25,000 | 1.11 [0.76, 1.62] | 0.580 |
| Household income: $50,000$–99,999 vs <$25,000 | 0.78 [0.55, 1.11] | 0.174 |
| Household income: ≥$100,000 vs$ <25,000 | 0.89 [0.61, 1.30] | 0.550 |
| Household income: Missing/Declined vs <$25,000 | 1.53 [0.98, 2.38] | 0.062 |
| Parental education: Some college / Associate vs Bachelor’s degree or higher | 1.10 [0.85, 1.43] | 0.470 |
| Parental education: High school or less vs Bachelor’s degree or higher | 1.09 [0.78, 1.54] | 0.607 |

[1] Multinomial logistic regression was used to examine associations between baseline parent psychopathology and child longitudinal trajectory type. Models were refit using alternative reference groups to obtain all pairwise trajectory comparisons. Parent ASR internalizing and externalizing scores were standardized, so odds ratios reflect the change in odds of membership in the first trajectory group relative to the second per 1-SD increase in parent symptoms. Models adjusted for child sex, child age at baseline, household income, and parental education. Estimates for the parental education Missing/Declined category were not displayed because of sparse cell counts across trajectory groups.

**Table S4. Aim 1 sensitivity analysis: retaining only one sibling per family to determine whether class trajectory membership would change.**

| Outcome | Full sample (N = 11,142) | One participant per family (N = 9,371) | Difference |
|---|---|---|---|
| **Trajectory type** | | | |
| Stable | 76.1% | 74.8% | -1.3 |
| Improved | 9.9% | 10.9% | +1.0 |
| Worsened | 8.1% | 7.9% | -0.2 |
| Fluctuate | 5.9% | 6.3% | +0.4 |
| **Transition probabilities** | | | |
| Mean absolute difference | — | — | 0.006 |
| Maximum absolute difference | — | — | 0.028 |

For trajectory types, differences in the number of participants who endorse each trajectory class are expressed as a percentage and calculated as the one-participant-per-family estimate minus the full-sample estimate. Transition probability differences represent absolute differences between corresponding model-estimated transition probabilities.

**Table S5. Aim 2 sensitivity analysis: retaining only one sibling per family to determine whether class CLPM-derived parent-child associations would differ.**

| Pathway | Full sample Standardized β | One participant per family Standardized β | Δβ | Full sample p-value | One participant per family p-value |
|---|---|---|---|---|---|
| **Child internalizing** | | | | | |
| Parent internalizing baseline → Child internalizing 2-year | 0.120 | 0.124 | +0.004 | <.001 | <.001 |
| Parent externalizing baseline → Child internalizing 2-year | 0.036 | 0.041 | +0.005 | 0.021 | 0.020 |
| Parent internalizing 2-year → Child internalizing 4-year | 0.117 | 0.118 | +0.001 | <.001 | <.001 |
| Parent externalizing 2-year → Child internalizing 4-year | 0.029 | 0.027 | -0.002 | 0.046 | 0.083 |
| Child internalizing baseline → Parent internalizing 2-year | 0.044 | 0.041 | -0.003 | <.001 | <.001 |
| Child internalizing 2-year → Parent internalizing 4-year | 0.048 | 0.050 | +0.002 | <.001 | <.001 |
| Child internalizing baseline → Parent externalizing 2-year | 0.035 | 0.035 | -0.000 | 0.003 | 0.005 |
| Child internalizing 2-year → Parent externalizing 4-year | 0.032 | 0.035 | +0.003 | 0.006 | 0.004 |
| **Child externalizing** | | | | | |

| | | | | | |
|---|---|---|---|---|---|
| Parent internalizing baseline → Child externalizing 2-year | 0.043 | 0.040 | -0.003 | 0.009 | 0.037 |
| Parent externalizing baseline → Child externalizing 2-year | 0.043 | 0.047 | +0.004 | 0.006 | 0.023 |
| Parent internalizing 2-year → Child externalizing 4-year | 0.031 | 0.038 | +0.008 | 0.080 | 0.030 |
| Parent externalizing 2-year → Child externalizing 4-year | 0.070 | 0.068 | -0.003 | <.001 | <.001 |
| Child externalizing baseline → Parent internalizing 2-year | 0.005 | 0.004 | -0.001 | 0.639 | 0.704 |
| Child externalizing 2-year → Parent internalizing 4-year | 0.012 | 0.018 | +0.006 | 0.300 | 0.241 |
| Child externalizing baseline → Parent externalizing 2-year | 0.023 | 0.022 | -0.001 | 0.077 | 0.112 |
| Child externalizing 2-year → Parent externalizing 4-year | 0.025 | 0.025 | -0.000 | 0.056 | 0.099 |

Standardized cross-lagged estimates are shown for the primary full-sample models and the sensitivity models restricted to one participant per family. Δβ is calculated as the one-participant-per-family standardized estimate minus the full-sample standardized estimate.

**Table S6. Comparison of CLPM and RI-CLPM cross-lagged path estimates**

| Model | Pathway | CLPM β | RI-CLPM β | Δβ[1] | CLPM p-value | RI-CLPM p-value |
|---|---|---|---|---|---|---|
| Child internalizing | Parent internalizing, baseline → Child internalizing, 2-year | 0.120 | 0.051 | -0.069 | <0.001 | 0.006 |
| Child internalizing | Parent internalizing, 2-year → Child internalizing, 4-year | 0.117 | 0.036 | -0.081 | <0.001 | 0.023 |
| Child internalizing | Child internalizing, baseline → Parent internalizing, 2-year | 0.044 | 0.047 | +0.003 | <0.001 | <0.001 |
| Child internalizing | Child internalizing, 2-year → Parent internalizing, 4-year | 0.048 | 0.011 | -0.037 | <0.001 | 0.543 |
| Child externalizing | Parent externalizing, baseline → Child externalizing, 2-year | 0.043 | 0.038 | -0.004 | 0.006 | 0.015 |
| Child externalizing | Parent externalizing, 2-year → Child externalizing, 4-year | 0.070 | 0.013 | -0.057 | <0.001 | 0.539 |
| Child externalizing | Child externalizing, baseline → Parent externalizing, 2-year | 0.023 | 0.056 | +0.034 | 0.077 | <0.001 |
| Child externalizing | Child externalizing, 2-year → Parent externalizing, 4-year | 0.025 | 0.016 | -0.009 | 0.056 | 0.451 |

[1] Δβ represents the RI-CLPM standardized estimate minus the corresponding CLPM standardized estimate.

**Table S7. Difference in Standardized Beta Estimates based on whether the same caregiver filled out the ASR/CBCL at each timepoint (Stable Group β; 92.5% of sample) or whether there is variation in the caregiver who filled out the ASR/CBCL across timepoints (Primary Group β; full sample)**

| Caregiver group | Pathway | Primary Group β | Stable Group β | Difference |
|---|---|---|---|---|
| **Child externalizing** | | | | |
| Father caregiver | child_ext_00A -> parent_ext_02A | 0.034 | 0.036 | 0.003 |
| Father caregiver | child_ext_00A -> parent_int_02A | 0.007 | 0.036 | 0.029 |
| Father caregiver | child_ext_02A -> parent_ext_04A | 0.001 | -0.022 | -0.024 |
| Father caregiver | child_ext_02A -> parent_int_04A | 0.002 | -0.017 | -0.019 |
| Father caregiver | parent_ext_00A -> child_ext_02A | -0.004 | 0.046 | 0.050 |
| Father caregiver | parent_ext_02A -> child_ext_04A | -0.000 | -0.012 | -0.012 |
| Father caregiver | parent_int_00A -> child_ext_02A | -0.001 | -0.064 | -0.063 |
| Father caregiver | parent_int_02A -> child_ext_04A | -0.010 | -0.027 | -0.016 |
| Mother caregiver | child_ext_00A -> parent_ext_02A | 0.014 | 0.008 | -0.006 |
| Mother caregiver | child_ext_00A -> parent_int_02A | -0.009 | -0.013 | -0.003 |
| Mother caregiver | child_ext_02A -> parent_ext_04A | 0.032 | 0.033 | 0.001 |
| Mother caregiver | child_ext_02A -> parent_int_04A | 0.020 | 0.021 | 0.001 |
| Mother caregiver | parent_ext_00A -> child_ext_02A | 0.050 | 0.059 | 0.009 |

| | | | | |
|---|---|---|---|---|
| Mother caregiver | parent_ext_02A -> child_ext_04A | 0.088 | 0.098 | 0.010 |
| Mother caregiver | parent_int_00A -> child_ext_02A | 0.048 | 0.043 | -0.004 |
| Mother caregiver | parent_int_02A -> child_ext_04A | 0.031 | 0.030 | -0.001 |
| **Child internalizing** | | | | |
| Father caregiver | child_int_00A -> parent_ext_02A | 0.033 | 0.072 | 0.040 |
| Father caregiver | child_int_00A -> parent_int_02A | 0.042 | 0.087 | 0.045 |
| Father caregiver | child_int_02A -> parent_ext_04A | -0.008 | -0.015 | -0.007 |
| Father caregiver | child_int_02A -> parent_int_04A | 0.023 | 0.024 | 0.001 |
| Father caregiver | parent_ext_00A -> child_int_02A | 0.056 | 0.115 | 0.059 |
| Father caregiver | parent_ext_02A -> child_int_04A | -0.015 | -0.081 | -0.066 |
| Father caregiver | parent_int_00A -> child_int_02A | 0.057 | -0.025 | -0.082 |
| Father caregiver | parent_int_02A -> child_int_04A | 0.079 | 0.153 | 0.074 |
| Mother caregiver | child_int_00A -> parent_ext_02A | 0.028 | 0.026 | -0.002 |
| Mother caregiver | child_int_00A -> parent_int_02A | 0.036 | 0.036 | 0.001 |
| Mother caregiver | child_int_02A -> parent_ext_04A | 0.034 | 0.032 | -0.003 |
| Mother caregiver | child_int_02A -> parent_int_04A | 0.049 | 0.045 | -0.004 |

| | | | | |
|---|---|---|---|---|
| Mother caregiver | parent_ext_00A -> child_int_02A | 0.031 | 0.037 | 0.006 |
| Mother caregiver | parent_ext_02A -> child_int_04A | 0.037 | 0.041 | 0.004 |
| Mother caregiver | parent_int_00A -> child_int_02A | 0.128 | 0.128 | 0.000 |
| Mother caregiver | parent_int_02A -> child_int_04A | 0.112 | 0.110 | -0.002 |

Results from the stable caregiver sensitivity analysis were highly similar to the primary analyses for mother-child models. Some attenuation and variability were observed in father-child models, likely reflecting the smaller sample size after restricting to participants with stable caregiver reporters. Overall, the direction and pattern of findings remained largely consistent, suggesting that the results of the main Aim 2 analysis are not being driven by variation in caregiver administering the ASR and CBCL.

**Table S8. Inverse probability weighted sensitivity analysis of parent-child cross-lagged paths**

| Caregiver group | Pathway | Unweighted standardized β[1] | Unweighted p-value | IPW standardized β[1] | IPW p-value | Difference in standardized β[1] | Unweighted path-specific Z-score[2] | IPW path-specific Z-score[2] |
|---|---|---|---|---|---|---|---|---|
| **Child externalizing** | | | | | | | | |
| Father caregiver | child_ext_00A → parent_ext_02A | 0.034 | 0.466 | 0.026 | 0.602 | -0.008 | 0.199 | 0.367 |
| Father caregiver | child_ext_00A → parent_int_02A | 0.007 | 0.832 | -0.001 | 0.967 | -0.008 | 0.735 | 0.784 |
| Father caregiver | child_ext_02A → parent_ext_04A | 0.001 | 0.981 | -0.025 | 0.599 | -0.026 | 0.988 | 0.513 |
| Father caregiver | child_ext_02A → parent_int_04A | 0.002 | 0.961 | -0.008 | 0.869 | -0.010 | 0.974 | 0.911 |
| Father caregiver | parent_ext_00A → child_ext_02A | -0.004 | 0.932 | 0.011 | 0.842 | 0.015 | 0.906 | 0.953 |
| Father caregiver | parent_ext_02A → child_ext_04A | -0.000 | 0.996 | -0.015 | 0.740 | -0.015 | 0.892 | 0.746 |
| Father caregiver | parent_int_00A → child_ext_02A | -0.001 | 0.976 | -0.003 | 0.956 | -0.001 | 0.870 | 0.869 |
| Father caregiver | parent_int_02A → child_ext_04A | -0.010 | 0.822 | -0.003 | 0.955 | 0.008 | 0.821 | 0.941 |
| Mother caregiver | child_ext_00A → parent_ext_02A | 0.014 | 0.296 | 0.014 | 0.335 | -0.001 | 1.133 | 1.292 |

| | | | | | | | | |
|---|---|---|---|---|---|---|---|---|
| Mother caregiver | child_ext_00A → parent_int_02A | -0.009 | 0.430 | -0.010 | 0.423 | -0.000 | 5.533 | 5.681 |
| Mother caregiver | child_ext_02A → parent_ext_04A | 0.032 | 0.029 | 0.032 | 0.025 | 0.000 | 0.740 | 0.749 |
| Mother caregiver | child_ext_02A → parent_int_04A | 0.020 | 0.151 | 0.020 | 0.143 | 0.001 | 0.463 | 0.368 |
| Mother caregiver | parent_ext_00A → child_ext_02A | 0.050 | 0.011 | 0.050 | 0.009 | 0.000 | 0.160 | 0.098 |
| Mother caregiver | parent_ext_02A → child_ext_04A | 0.088 | <0.001 | 0.088 | <0.001 | 0.000 | 3.384 | 3.520 |
| Mother caregiver | parent_int_00A → child_ext_02A | 0.048 | 0.009 | 0.046 | 0.009 | -0.001 | 0.278 | 0.333 |
| Mother caregiver | parent_int_02A → child_ext_04A | 0.031 | 0.098 | 0.030 | 0.112 | -0.001 | 0.784 | 0.767 |
| **Child internalizing** | | | | | | | | |
| Father caregiver | child_int_00A → parent_ext_02A | 0.033 | 0.417 | 0.021 | 0.661 | -0.012 | 0.873 | 0.854 |
| Father caregiver | child_int_00A → parent_int_02A | 0.042 | 0.235 | 0.036 | 0.321 | -0.006 | 0.716 | 0.738 |
| Father caregiver | child_int_02A → parent_ext_04A | -0.008 | 0.855 | -0.032 | 0.471 | -0.024 | 0.404 | 1.215 |
| Father caregiver | child_int_02A → parent_int_04A | 0.023 | 0.585 | 0.006 | 0.889 | -0.016 | 0.788 | 0.553 |

| | | | | | | | | |
|---|---|---|---|---|---|---|---|---|
| Father caregiver | parent_ext_00A → child_int_02A | 0.056 | 0.236 | 0.077 | 0.186 | 0.021 | 0.558 | 0.049 |
| Father caregiver | parent_ext_02A → child_int_04A | -0.015 | 0.718 | -0.030 | 0.514 | -0.014 | 0.434 | 0.616 |
| Father caregiver | parent_int_00A → child_int_02A | 0.057 | 0.208 | 0.054 | 0.287 | -0.003 | 0.558 | 0.585 |
| Father caregiver | parent_int_02A → child_int_04A | 0.079 | 0.075 | 0.096 | 0.045 | 0.016 | 0.168 | 0.693 |
| Mother caregiver | child_int_00A → parent_ext_02A | 0.028 | 0.022 | 0.027 | 0.031 | -0.001 | 3.901 | 4.263 |
| Mother caregiver | child_int_00A → parent_int_02A | 0.036 | 0.002 | 0.035 | 0.003 | -0.001 | 2.356 | 2.641 |
| Mother caregiver | child_int_02A → parent_ext_04A | 0.034 | 0.006 | 0.035 | 0.005 | 0.000 | 2.648 | 2.624 |
| Mother caregiver | child_int_02A → parent_int_04A | 0.049 | <0.001 | 0.049 | <0.001 | -0.000 | 0.314 | 0.288 |
| Mother caregiver | parent_ext_00A → child_int_02A | 0.031 | 0.073 | 0.031 | 0.073 | 0.000 | 1.035 | 1.037 |
| Mother caregiver | parent_ext_02A → child_int_04A | 0.037 | 0.022 | 0.039 | 0.021 | 0.001 | 0.268 | 0.129 |
| Mother caregiver | parent_int_00A → child_int_02A | 0.128 | <0.001 | 0.128 | <0.001 | 0.000 | 8.652 | 8.920 |
| Mother caregiver | parent_int_02A → child_int_04A | 0.112 | <0.001 | 0.112 | <0.001 | -0.001 | 5.832 | 5.821 |

| |
|---|
| [1] Standardized estimates are std.all estimates from the unweighted and IPW-weighted CLPMs. Difference was calculated as IPW standardized β minus unweighted standardized β. |
| [2] The Z-scores in the table represent the contribution of the respective path standardized estimate to the model-wide Pearson correlation within each iteration. Within each of the 5000 iterations, we split the sample into two split-halves (A and B), and calculated a model-wide Pearson correlation coefficient encompassing the correlations across all the respective standardized beta estimates between the two split-halves. We then calculated the contribution of each path in this Pearson correlation coefficient (1 coefficient for each iteration). We quantified the absolute mean contribution divided by the standard deviation across all iterations to calculate the Z-scores provided in this table. Larger Z-scores indicates a more reproducible contribution of that pathway to the model-wide split-half Pearson correlation. These values should not be interpreted as pathway-specific Pearson correlations, effect sizes, or tests of statistical significance for individual paths. |

**Table S9. Probability of endorsing a T-score>65 across CBCL domains at baseline by latent class**

| Symptom | Low symptom group | Moderate symptoms | High symptom group |
|---|---|---|---|
| Aggressive Behavior | 0.003 | 0.094 | 0.679 |
| Anxious/Depressed | 0.006 | 0.197 | 0.605 |
| Attention Problems | 0.006 | 0.164 | 0.661 |
| Rule-Breaking Behavior | 0.004 | 0.079 | 0.457 |
| Social Problems | 0.001 | 0.055 | 0.522 |
| Somatic Complaints | 0.020 | 0.219 | 0.421 |
| Thought Problems | 0.005 | 0.172 | 0.718 |
| Withdrawn/Depressed | 0.007 | 0.199 | 0.527 |

**Table S10. Model-estimated latent class transition probabilities (Aim 1)**

| Current class | Low | Moderate | High |
|---|---|---|---|
| **Baseline -> 1-year** | | | |
| Low | 0.96 | 0.03 | 0.00 |
| Moderate | 0.16 | 0.79 | 0.05 |
| High | 0.04 | 0.26 | 0.71 |
| **1-year -> 2-year** | | | |
| Low | 0.96 | 0.04 | 0.00 |
| Moderate | 0.15 | 0.77 | 0.08 |
| High | 0.03 | 0.26 | 0.71 |
| **2-year -> 3-year** | | | |
| Low | 0.97 | 0.03 | 0.00 |
| Moderate | 0.13 | 0.80 | 0.06 |
| High | 0.02 | 0.28 | 0.70 |
| **3-year -> 4-year** | | | |
| Low | 0.95 | 0.04 | 0.00 |
| Moderate | 0.15 | 0.75 | 0.10 |
| High | 0.04 | 0.22 | 0.74 |
| **4-year -> 5-year** | | | |
| Low | 0.97 | 0.02 | 0.00 |
| Moderate | 0.15 | 0.80 | 0.06 |
| High | 0.02 | 0.27 | 0.71 |

**Table S11. Parent-child cross-lagged panel models of internalizing and externalizing symptoms**

| Outcome | Pathway | Standardized β | 95% CI | P value |
|---|---|---|---|---|
| Child internalizing model | Parent internalizing baseline → Child internalizing 2-year | 0.120 | [0.090, 0.151] | <0.001 |
| Child internalizing model | Parent externalizing baseline → Child internalizing 2-year | 0.036 | [0.005, 0.067] | 0.021 |
| Child internalizing model | Parent internalizing 2-year → Child internalizing 4-year | 0.117 | [0.089, 0.145] | <0.001 |
| Child internalizing model | Parent externalizing 2-year → Child internalizing 4-year | 0.029 | [0.000, 0.058] | 0.046 |
| Child internalizing model | Child internalizing baseline → Parent internalizing 2-year | 0.044 | [0.022, 0.066] | <0.001 |
| Child internalizing model | Child internalizing 2-year → Parent internalizing 4-year | 0.048 | [0.026, 0.070] | <0.001 |
| Child internalizing model | Child internalizing baseline → Parent externalizing 2-year | 0.035 | [0.012, 0.059] | 0.003 |
| Child internalizing model | Child internalizing 2-year → Parent externalizing 4-year | 0.032 | [0.009, 0.054] | 0.006 |
| Child externalizing model | Parent internalizing baseline → Child externalizing 2-year | 0.043 | [0.011, 0.076] | 0.009 |
| Child externalizing model | Parent externalizing baseline → Child externalizing 2-year | 0.043 | [0.012, 0.073] | 0.006 |
| Child externalizing model | Parent internalizing 2-year → Child externalizing 4-year | 0.031 | [-0.004, 0.065] | 0.080 |
| Child externalizing model | Parent externalizing 2-year → Child externalizing 4-year | 0.070 | [0.031, 0.109] | <0.001 |

| | | | | |
|---|---|---|---|---|
| Child externalizing model | Child externalizing baseline → Parent internalizing 2-year | 0.005 | [-0.016, 0.026] | 0.639 |
| Child externalizing model | Child externalizing 2-year → Parent internalizing 4-year | 0.012 | [-0.011, 0.036] | 0.300 |
| Child externalizing model | Child externalizing baseline → Parent externalizing 2-year | 0.023 | [-0.002, 0.048] | 0.077 |
| Child externalizing model | Child externalizing 2-year → Parent externalizing 4-year | 0.025 | [-0.001, 0.051] | 0.056 |

**Table S12. Equality-constraint tests comparing parent-to-child and child-to-parent cross-lagged paths**

| Parent domain | Child domain | Time interval | Parent → child β | Child → parent β | Numerically larger path | $\chi^2$ | df | p-value |
|---|---|---|---|---|---|---|---|---|
| **Child internalizing model** | | | | | | | | |
| Internalizing | Internalizing | Baseline to 2-year | 0.120 | 0.044 | Parent -> child | 25.278 | 1 | <0.001 |
| Internalizing | Internalizing | 2-year to 4-year | 0.117 | 0.048 | Parent -> child | 19.575 | 1 | <0.001 |
| Externalizing | Internalizing | Baseline to 2-year | 0.036 | 0.035 | Parent -> child | 0.006 | 1 | 0.940 |
| Externalizing | Internalizing | 2-year to 4-year | 0.029 | 0.032 | Child -> parent | 0.032 | 1 | 0.858 |
| **Child externalizing model** | | | | | | | | |
| Internalizing | Externalizing | Baseline to 2-year | 0.043 | 0.005 | Parent -> child | 7.033 | 1 | 0.008 |

| | | | | | | | | |
|---|---|---|---|---|---|---|---|---|
| Internalizing | Externalizing | 2-year to 4-year | 0.031 | 0.012 | Parent -> child | 1.497 | 1 | 0.221 |
| Externalizing | Externalizing | Baseline to 2-year | 0.043 | 0.023 | Parent -> child | 1.816 | 1 | 0.178 |
| Externalizing | Externalizing | 2-year to 4-year | 0.070 | 0.025 | Parent -> child | 9.166 | 1 | 0.002 |

[1] Models were fit using standardized symptom variables so that directional parent-to-child and child-to-parent paths could be compared on a common scale. Equality constraints were tested using Wald tests in lavaan. The numerically larger path column indicates which direction had the larger absolute coefficient; statistical evidence for directional differences is shown by the Wald test p-value. Results confirm that parent-child relationships are stronger than child-parent relationships.

**Table S13. Comparing child sex and household income representation across psychopathology trajectory type**

| Demographic Variable of Interest | Trajectory type | n (%) | Chi-square p-value |
|---|---|---|---|
| **Child sex** | | | |
| Female | Stable | 4,002 (75.6%) | <0.001 |
| Female | Improved | 407 (7.7%) | |
| Female | Worsened | 578 (10.9%) | |
| Female | Fluctuate | 305 (5.8%) | |
| Male | Stable | 4,473 (76.5%) | |
| Male | Improved | 700 (12%) | |
| Male | Worsened | 330 (5.6%) | |
| Male | Fluctuate | 347 (5.9%) | |
| **Household income** | | | |
| <$50k | Stable | 2,066 (70.6%) | <0.001 |

| | | |
|---|---|---|
| <$50k | Improved | 392 (13.4%) |
| <$50k | Worsened | 247 (8.4%) |
| <$50k | Fluctuate | 223 (7.6%) |
| $50k–$99,999 | Stable | 2,204 (75.6%) |
| $50k–$99,999 | Improved | 288 (9.9%) |
| $50k–$99,999 | Worsened | 266 (9.1%) |
| $50k–$99,999 | Fluctuate | 157 (5.4%) |
| ≥$100k | Stable | 3,535 (80.3%) |
| ≥$100k | Improved | 323 (7.3%) |
| ≥$100k | Worsened | 339 (7.7%) |
| ≥$100k | Fluctuate | 206 (4.7%) |

[1] Trajectory types were derived from posterior class assignments across six waves. Income was grouped into three categories: <$50k, $50k–$99,999, and ≥$100k. Participants with missing or declined income were excluded from income-stratified summaries.

**Table S14. Comparing child sex and household income representation across baseline classes**

| Demographic Variable of Interest | Baseline class | n (%) | Chi-square p-value |
|---|---|---|---|
| **Child sex** | | | |
| Female | Low | 4,164 (78.7%) | <0.001 |
| Female | Moderate | 913 (17.3%) | |
| Female | High | 215 (4.1%) | |
| Male | Low | 4,391 (75.1%) | |
| Male | Moderate | 1,092 (18.7%) | |

| | | | |
|---|---|---|---|
| Male | High | 367 (6.3%) | |
| **Household income** | | | |
| <$50k | Low | 2,055 (70.2%) | <0.001 |
| <$50k | Moderate | 612 (20.9%) | |
| <$50k | High | 261 (8.9%) | |
| $50k–$99,999 | Low | 2,226 (76.4%) | |
| $50k–$99,999 | Moderate | 545 (18.7%) | |
| $50k–$99,999 | High | 144 (4.9%) | |
| ≥$100k | Low | 3,599 (81.7%) | |
| ≥$100k | Moderate | 687 (15.6%) | |
| ≥$100k | High | 117 (2.7%) | |

[1] Baseline class was assigned using the highest posterior class probability at baseline. Chi-square p-values compare the distribution of baseline latent classes across strata.

### 3. Figures

**Figure S1. Consort diagram detailing exclusion criteria for Aim 1 and 2**

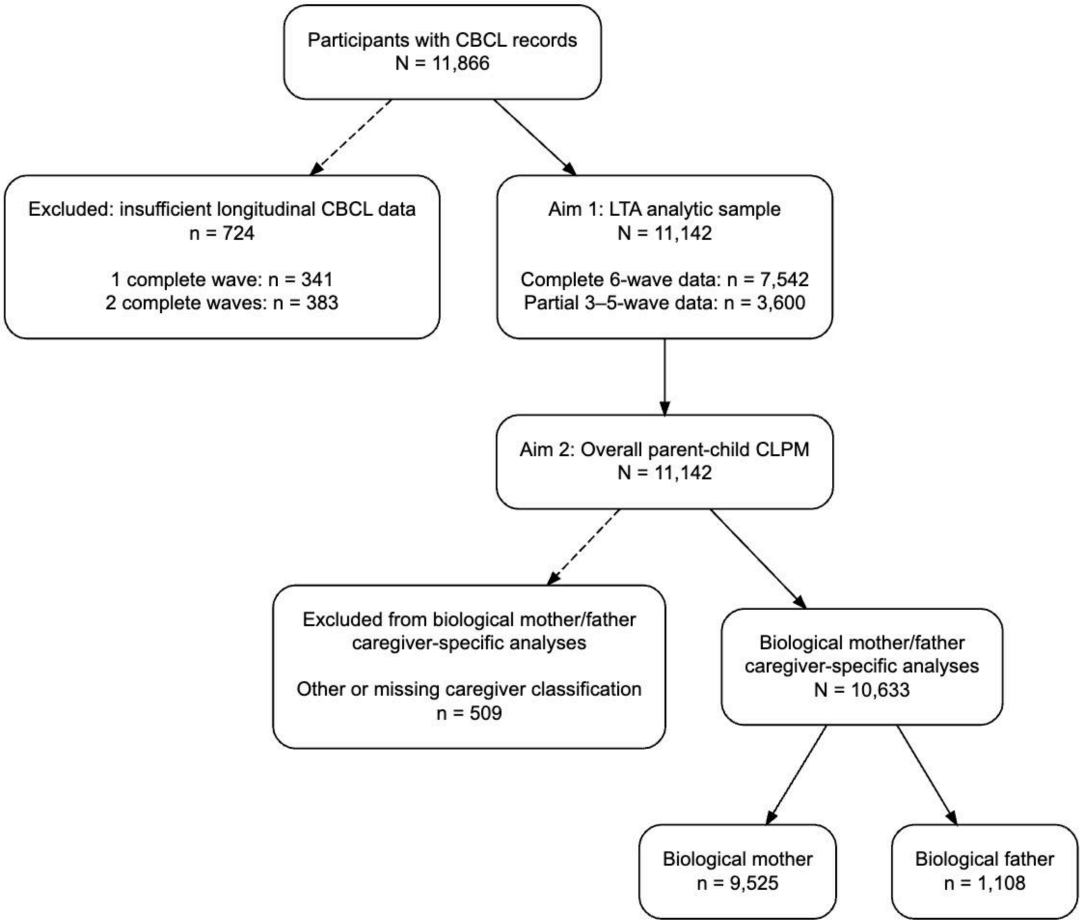

**Figure S2. Zero-order parent-child correlations of psychopathology subscale scores across sex.**

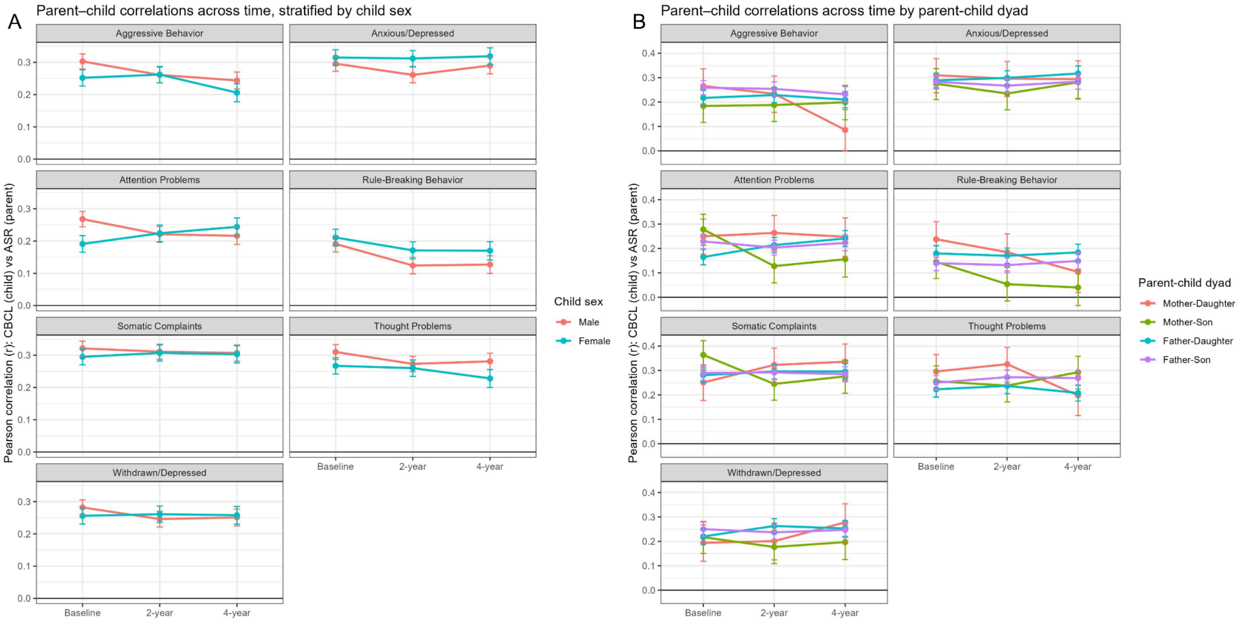

Note: Parents self-reported their symptoms using the Adult Rating Scale (ASR) and on their child's behaviors using the Child Behavioral Checklist (CBCL). No covariates were used in these correlations. (A) depicts parent (mother or father)-child correlations stratified by child sex. (B) depicts the parent-child correlations stratified by both parent and child sex.
